# Pulmonary Vein Isolation Versus Left Atrial Posterior Wall Isolation with the Cryoballoon Using the Cross-Over Technique in Persistent Atrial Fibrillation

**DOI:** 10.1101/2024.03.04.24303746

**Authors:** Fuminori Odagiri, Takashi Tokano, Tetsuro Miyazaki, Koji Hirabayashi, Kai Ishi, Hiroshi Abe, Sayaki Ishiwata, Midori Kakihara, Masaaki Maki, Hiroki Matsumoto, Ryosuke Shimai, Tadao Aikawa, Shintaro Takano, Yuki Kimura, Shunsuke Kuroda, Hiroyuki Isogai, Dai Ozaki, Tomoyuki Shiozawa, Yuki Yasuda, Kiyoshi Takasu, Kenichi Iijima, Kazuhisa Takamura, Tomomi Matsubara, Haruna Tabuchi, Hidemori Hayashi, Ken Yokoyama, Gaku Sekita, Masataka Sumiyoshi, Yuji Nakazato, Tohru Minamino

## Abstract

**Background:** The cryoballoon (CB) has recently been used for performing pulmonary vein isolation (PVI) with concomitant left atrial posterior wall isolation (LAPWI). However, successful LAPWI using only the CB is technically challenging. This study aimed to evaluate the feasibility, safety, and efficacy of the cross-over technique, wherein an overlapped ablation is performed by placing the CB from both left and right directions in contact with the roof and bottom of the left atrium (LA).

**Methods:** This was a single-center, retrospective, observational study of 194 consecutive patients with persistent atrial fibrillation (PerAF) who underwent a first-time procedure of PVI + PWI (108 patients) or PVI-only (86 patients) using the CB between April 2021 and December 2023. The cross-over technique was applied in all LAPWI using the CB.

**Results:** For ablation of the LA roof and bottom, respectively, a mean of 8.6 ± 1.0 (L→R 4.3 ± 1.1 and R→L 4.3 ± 1.1) and 9.1 ± 1.2 (L→R 4.5 ± 1.2 and R→L 4.6 ± 1.6) CB applications were delivered. LAPW was successfully isolated solely using CB in 107 patients (99.1%). The PVI + PWI group, compared to the PVI-only group, had significantly longer procedure time (180.0 ± 42.4 vs. 78.0 ± 22.1 min, *P* < 0.001). No severe adverse events were observed in either group. During a median follow-up of 19 months, 93.5% and 72.9% of the PVI + PWI and PVI-only groups, respectively, were free from all atrial tachyarrhythmia recurrence (*P* = 0.011). Cox regression analyses showed LA diameter and PVI + PWI were independent predictors of all atrial tachyarrhythmias recurrence.

**Conclusions:** LAPWI performed solely with the CB using the cross-over technique is feasibly, safe, and was independently associated with a significantly higher freedom from recurrence of atrial tachyarrhythmias compared with PVI alone on midterm follow-up in patients with PerAF.

**WHAT IS KNOWN?:**

- The cryoballoon (CB) has recently been used for performing pulmonary vein isolation (PVI) with concomitant left atrial posterior wall isolation (LAPWI). LAPWI using the CB was previously demonstrated to be superior to PVI alone for the treatment of persistent atrial fibrillation (PerAF).
- However, successful LAPWI using only the CB is technically challenging, and there is no established LAPWI technique or protocol using the CB.

**WHAT THE STUDY ADDS?:**

- LAPWI solely with the CB was successfully performed using the cross-over technique, wherein an overlapped ablation was performed placing the CB from both left to right and right to left directions in contact with the roof and bottom of the LA.
- This technique achieved a high rate of acute success (99.1%) and freedom from recurrence of atrial tachyarrhythmias (93.5%) on a midterm follow-up without increasing adverse events.
- The cross-over technique could be considered an effective ablation strategy for LAPWI, especially in difficult cases such as narrow and steeply angled LA roof and bottom.

## Introduction

Pulmonary vein isolation (PVI) is the primary strategy of catheter ablation for treating atrial fibrillation (AF)^1^, and it is particularly effective for paroxysmal AF^2^. However, PVI alone has limited efficacy in patients with persistent AF (PerAF)^3^, despite the use of numerous ablation techniques^4^. An optimal additional lesion set has not been established^5^.

The posterior wall of the left atrium (LA) is a target for catheter ablation^6^. The posterior wall of the LA is embryologically related to the pulmonary veins and is responsible for the initiation and maintenance of AF^7^. Richardson et al^8^. reported several reasons why the combination of PVI and posterior wall isolation (PWI) is an attractive lesion set. First, isolation of the entire posterior wall has been associated with improved outcomes^9^. Second, PWI creates a conduction block to prevent potential reentrant circuits. Third, PWI reinforces PVI by reducing the potential for small gaps in the posterior aspect of the PVI lesion set, leading to PV conduction recovery. Fourth, PWI debulks the potential substrate or trigger sites for AF initiation or perpetuation, since the posterior wall is histologically similar to PV tissue^10, 11^. Finally, PWI may more reliably lead to ablation of the ganglionated plexi.

The cryoballoon (CB) has recently been used for performing PVI with concomitant LAPWI, and several studies have shown that LAPWI using the CB is superior to PVI alone for the treatment of PerAF^12–16^. However, successful LAPWI using only the CB is technically challenging, with reported success rates ranging from 54% to 84%^17^. Moreover, no LAPWI technique and protocol using the CB has been formally established.

To address these issues for LAPWI using the CB, we devised the cross-over technique, wherein an overlapped ablation was performed placing the CB from both left to right and right to left directions in contact with the roof and bottom of the LA. This study aimed to evaluate the feasibility, safety, and efficacy of this approach for LAPWI compared with PVI alone in patients with PerAF.

## Methods

### Study population

This was a single-center retrospective observational study that included 194 consecutive patients with PerAF (lasting more than 7 days) who underwent a first-time procedure of PVI + PWI (108 patients) or PVI-only (86 patients) between April 2021 and December 2023. Additional LAPWI was performed at the discretion of the physician. Prior ablation of right atrial arrhythmia was permitted. The exclusion criteria included patients younger than 18 years, paroxysmal AF (terminated within 7 days), sustained atrial fibrillation lasting more than 3 years, prior left atrial ablation (catheter or surgically based), cerebral ischemic events, myocardial infarction, or unstable angina in the previous 2 months, intracavitary thrombus, significant valvular disease, corrected or uncorrected congenital heart disease, a left atrial diameter of 60 mm or greater, chronic kidney disease requiring dialysis, pregnancy, contraindications to general anesthesia, life expectancy less than 1 year, contraindications to oral anticoagulants (OACs), clinical follow-up less than 3 months, and inability to provide informed consent. The study protocol was conducted in accordance with the Declaration of Helsinki and approved by the local ethics committee of our institution. All patients provided written informed consent.

### Preprocedural management

Patient demographics were recorded at baseline. A transthoracic echocardiogram (TTE) was performed within 1 year before the procedure to assess structural heart disease. Antiarrhythmic drugs (AADs) were discontinued for longer than five half-lives before the procedure. Amiodarone was discontinued at least one month before the procedure when possible, at the attending cardiologist’s discretion. A transesophageal echocardiogram (TEE) was performed to exclude the presence of thrombi the day before the procedure. Contrast-enhanced multidetector cardiac computed tomography was performed before the procedure to assess detailed LA and PV anatomy. All patients received OACs for 3 weeks or longer before the procedure, and these drugs were not interrupted (warfarin and dabigatran) or were temporarily switched to dabigatran (rivaroxaban, apixaban, and edoxaban) for the procedure.

### Procedure, PVI

All procedures were performed under moderate sedation with dexmedetomidine, bolus injection of pentazocine and thiopental as needed, and a nasal mask for noninvasive mechanical ventilation. After obtaining vascular access, heparin (100 IU/kg body weight) was administered. Two transseptal catheters are necessary for mapping and catheter ablation in LAPWI. Accordingly, a single (PVI-only) or double (PVI + PWI) transseptal puncture was performed as needed with an RF needle (Baylis Medical, Inc, Montreal, Canada) guided by fluoroscopy and intracardiac echocardiography (ViewFlex, Abbott, St. Paul, MN, USA). After obtaining LA access, the activated clotting time was maintained at ≥ 300 seconds by heparin infusion.

In all study patients, a fourth-generation 28-mm CB catheter (Arctic Front Advance PRO; Medtronic), was introduced into the LA through a steerable sheath (FlexCath Advance Medtronic Inc, 15Fr), and an inner lumen mapping catheter (Achieve, Medtronic Inc, Minneapolis, MN, USA) was positioned in each PV ostium. The CB was advanced, inflated, and positioned at the ostium of each PV. After achieving optimal PV occlusion, as assessed by venous angiography, 1–2 CB applications were delivered for 180–240 s to each PV based on previous reports^18, 19^. Regarding the ablation sequence, the left superior PV (LSPV) was treated first, followed by the left inferior PV (LIPV), the right superior PV (RSPV), and then by the right inferior PV (RIPV) (Figure 1).

**Figure 1.**
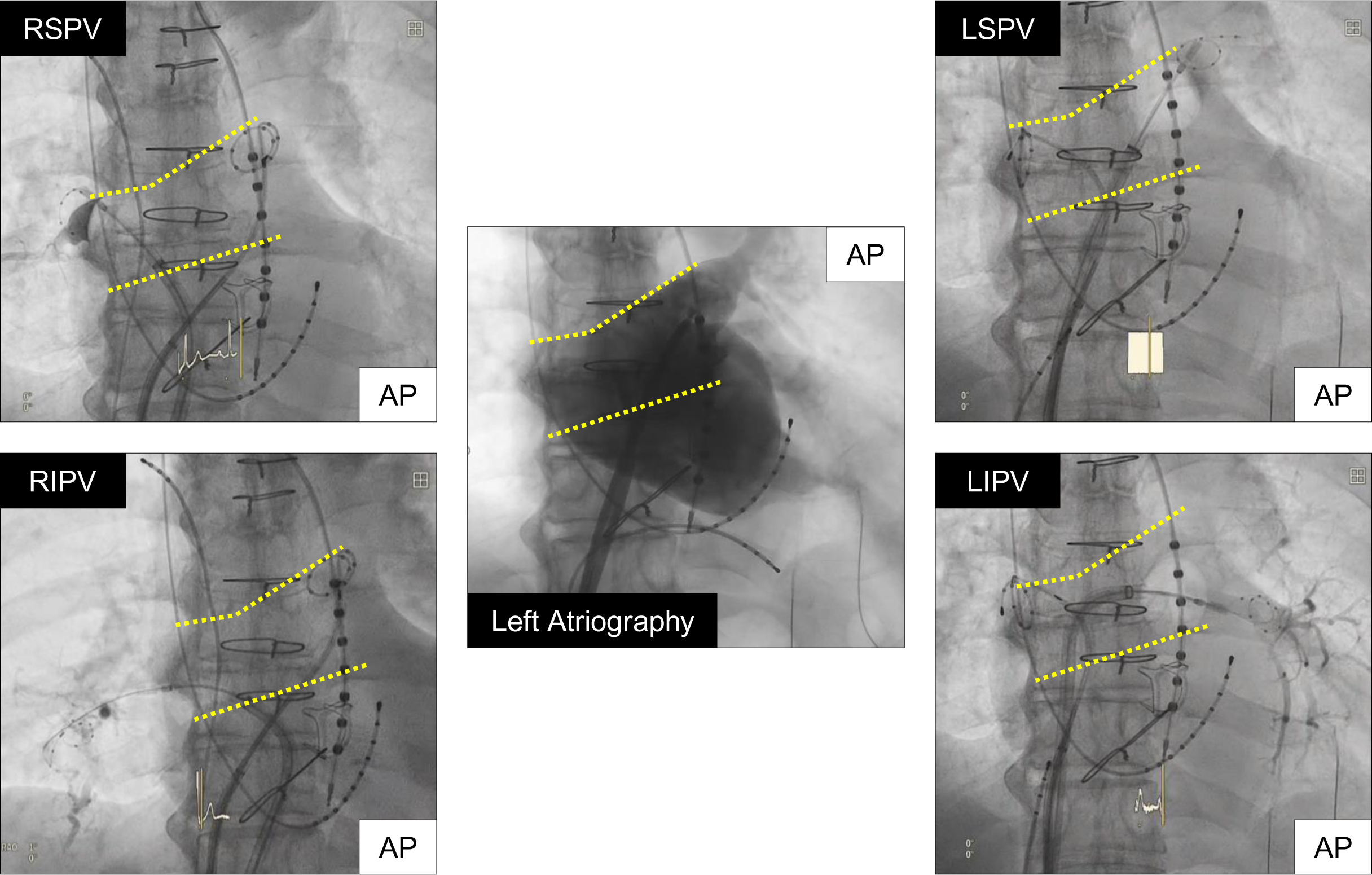
Representative AP fluoroscopic images of the left atriography and the CB position of during PVI. The LA roof and bottom were indicated by the yellow dotted lines. AP, anteroposterior; CB, cryoballoon; PVI, pulmonary vein isolation; LA, left atrial; LSPV, left superior pulmonary vein; LIPV, left inferior pulmonary vein; RSPV, right superior pulmonary vein; RIPV, right inferior pulmonary vein.

To avoid esophageal damage and gastric hypomotility (GH), the luminal esophageal temperature (LET) was monitored throughout ablation. CB applications were interrupted in case of LET ≤ 18℃. To prevent phrenic nerve (PN) injury, the diaphragmatic compound motor action potentials (CMAP) were monitored during PN pacing from an electrode in the right subclavian vein or superior vena cava during CB application of the right PVs. The CB application was immediately terminated if the maximum voltage of the CMAP decreased by 30% from baseline or if the diaphragmatic excursions weakened. Successful PVI was defined as a bidirectional block between PV and LA using an inner lumen mapping catheter (PVI-only) or a circular mapping catheter (Abbott Laboratories, Chicago, IL, USA) (PVI + PWI). No additional CB applications were delivered after PVI. If a successful PVI was not achieved, additional touch-up ablation was performed using an irrigated radiofrequency (RF) catheter (FlexAbility^TM^, Abbott, Chicago, IL, USA).

### Procedure, LAPWI

Once PVI was completed, CB was used to perform LAPWI. When LA roof area ablation was performed, the Achieve catheter was placed deeply in the LSPV to stabilize the CB, which was shifted clockwise along the LA roof by adjusting the bending of the steerable sheath to apply pressure. The LA roof area was divided into 4 segments from left to right guided by fluoroscopic images in the anteroposterior (AP) view, L1, L2, L3, and L4, toward the right superior PV (Figure 2). The CB was positioned such that each location overlapped with one-third of the previous site. A single 120-s CB application was delivered at each site.

**Figure 2.**
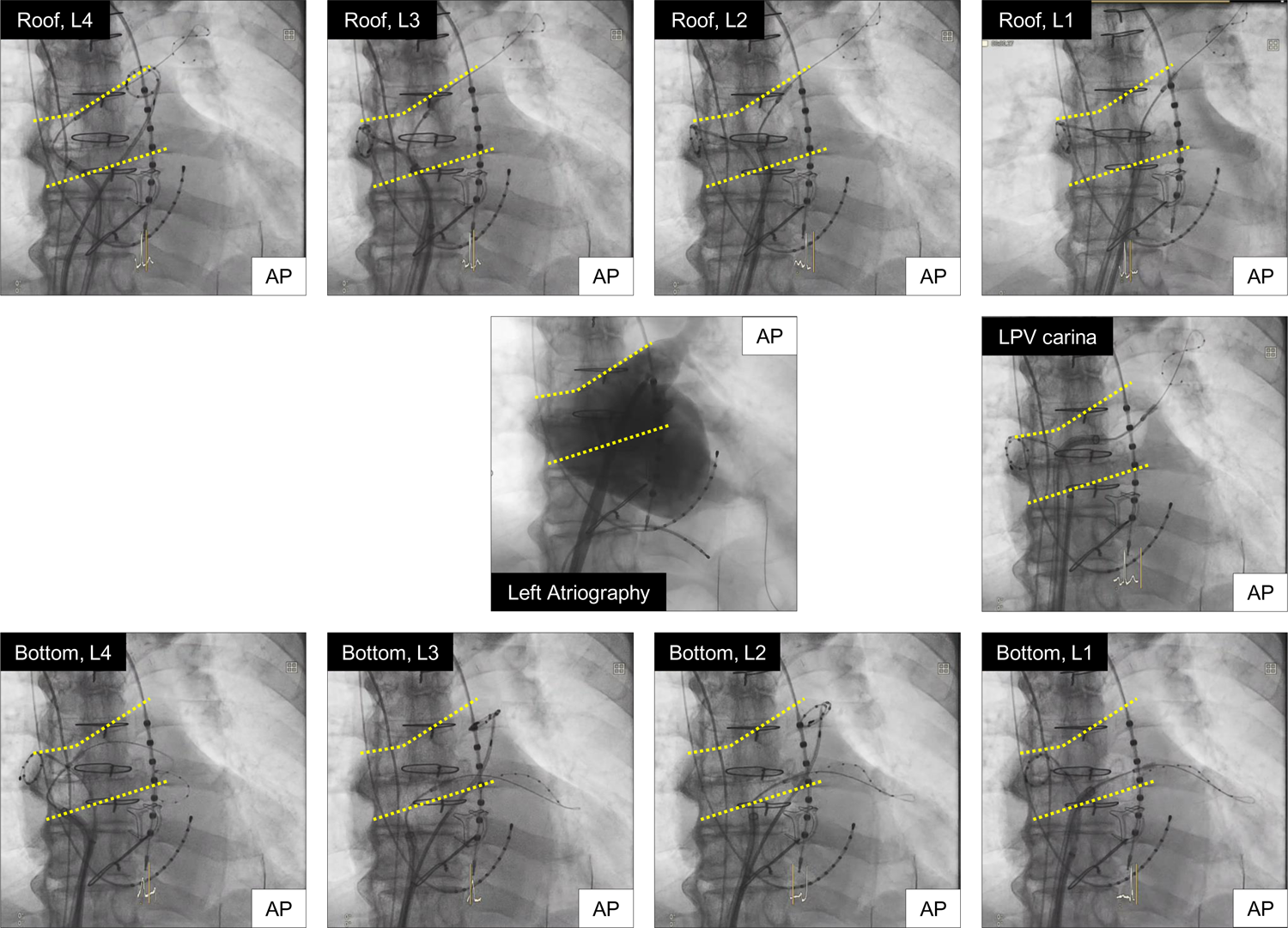
Representative AP fluoroscopic images of the left atriography, LA roof area, LA bottom area, and LPV carina ablation. The LA roof and bottom were divided into 4 segments from left to right guided by fluoroscopic images in the AP view, L1, L2, L3, and L4, toward the right side superior and inferior PV, respectively. The LA roof and bottom were indicated by the yellow dotted lines. AP, anteroposterior; LA, left atrial; LPV, left pulmonary vein; PV, pulmonary vein.

When LA bottom area ablation was performed, the Achieve catheter was deeply placed in the LIPV, and the CB was shifted clockwise along the LA bottom by adjusting the bending of the steerable sheath to apply pressure. The LA bottom area was also divided into 4 segments from left to right guided by fluoroscopic images in the AP view, L1, L2, L3, and L4, toward the right inferior PV (Figure 2). The left pulmonary vein (LPV) carina was routinely ablated with a single 180-s CB application by anchoring the achieve catheter in the LSPV, bending the steerable sheath (similar to when ablating the LIPV), and contacting the CB in a direction from the LSPV to the LIPV (Figure 2).

After the LAPW was ablated from left to right, the Achieve catheter was placed deeply in the RSPV, and the CB was shifted counterclockwise along the LA roof by adjusting the bending of the steerable sheath to apply pressure. The LA roof area was divided into 4 segments from right to left guided by fluoroscopic images in the AP view, R1, R2, R3, and R4, toward the left superior PV (Figure 3). CB ablation of the LA bottom area and right pulmonary vein (RPV) carina were performed in the same manner from the right side with the Achieve catheter positioned in the inferior and superior PV, respectively (Figure 3); this was the cross-over technique.

**Figure 3.**
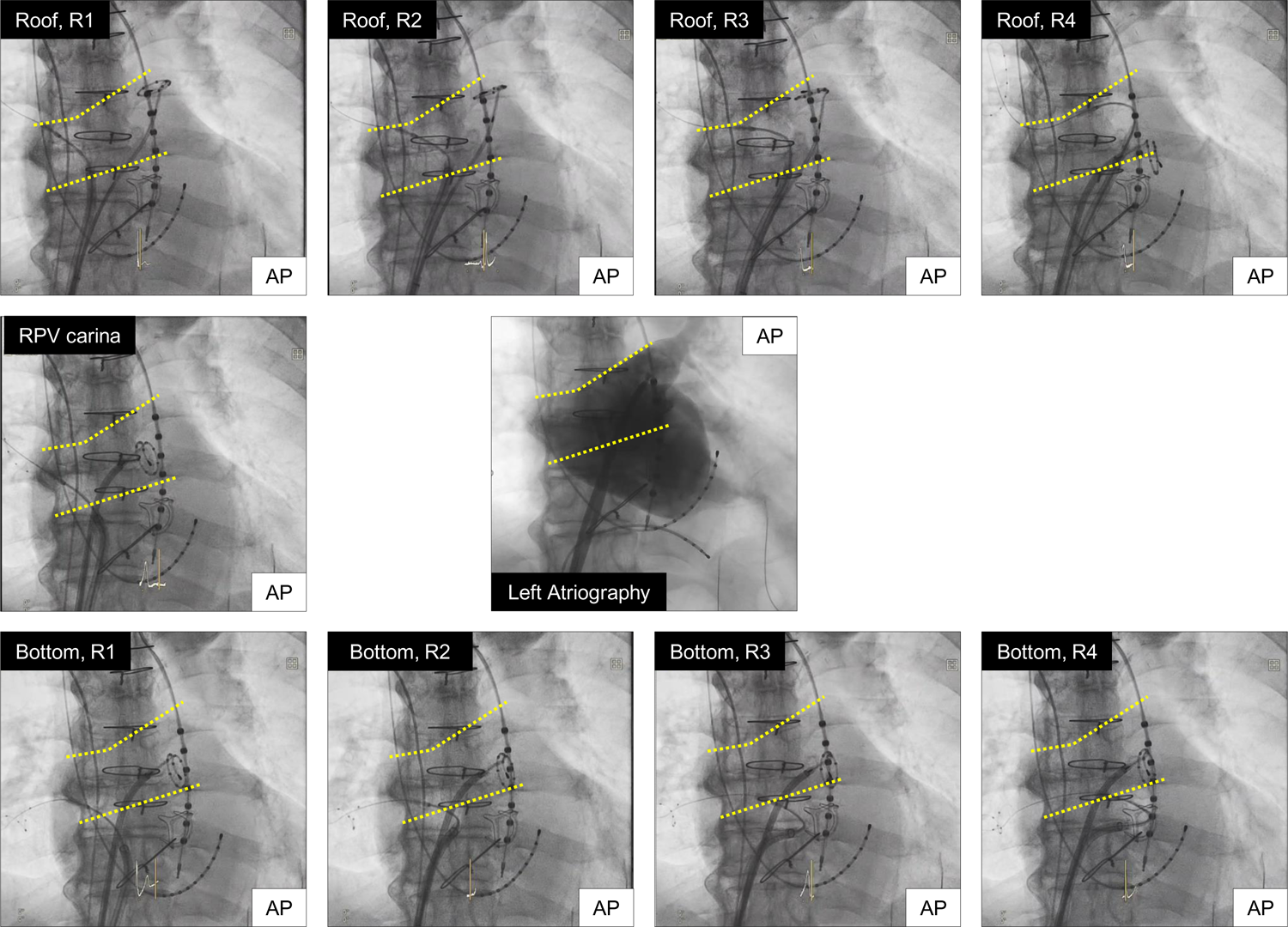
Representative AP fluoroscopic images of the left atriography, LA roof area, LA bottom area, and RPV carina ablation. The LA roof and bottom were divided into 4 segments from right to left guided by fluoroscopic images in the AP view, R1, R2, R3, and R4, toward the left side superior and inferior PV, respectively. The LA roof and bottom were indicated by the yellow dotted lines. AP, anteroposterior; LA, left atrial; RPV, right pulmonary vein; PV, pulmonary vein.

The LET was monitored throughout ablation, and temperatures ≤ 18℃ were avoided. CB applications were resumed once the LET returned to baseline. To prevent PN injury, CMAP was monitored during PN pacing from an electrode in the right subclavian vein or superior vena cava during CB ablation of the roof in R1, bottom in R1, and RPV carina.

Once completed, LAPWI was confirmed by creating voltage maps and using high output pacing (10V) with a circular mapping catheter to test for entrance or exit block (Figure 4-1 and 2). Whenever LAPWI could not be achieved between 1–2 additional applications using the CB alone at each site, additional touch-up ablation was performed using an irrigated RF catheter (FlexAbility^TM^, Abbott, Chicago, IL, USA). All procedures were performed by a single operator (F.O) skilled in this technique.

**Figure 4-1 and 2.**
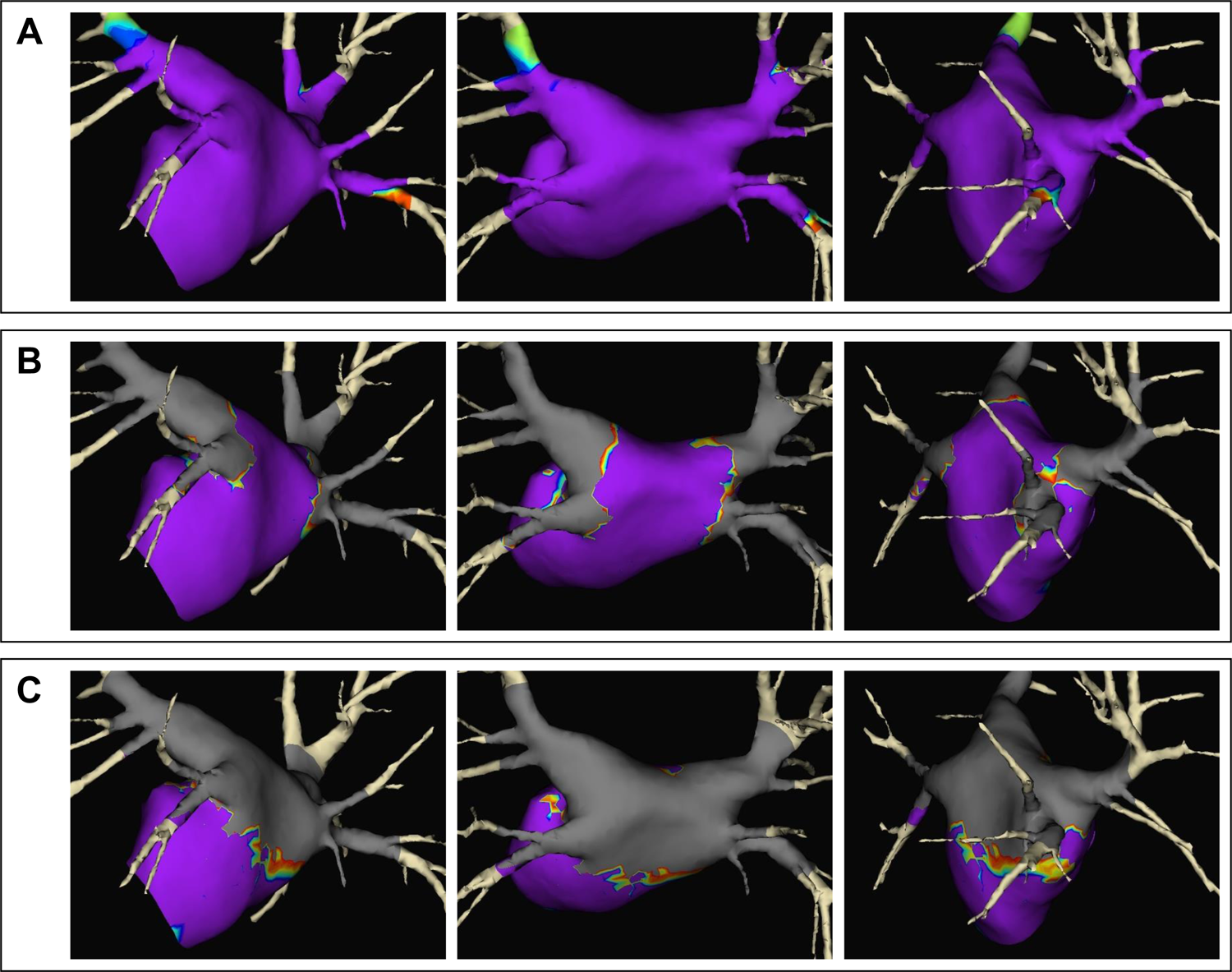
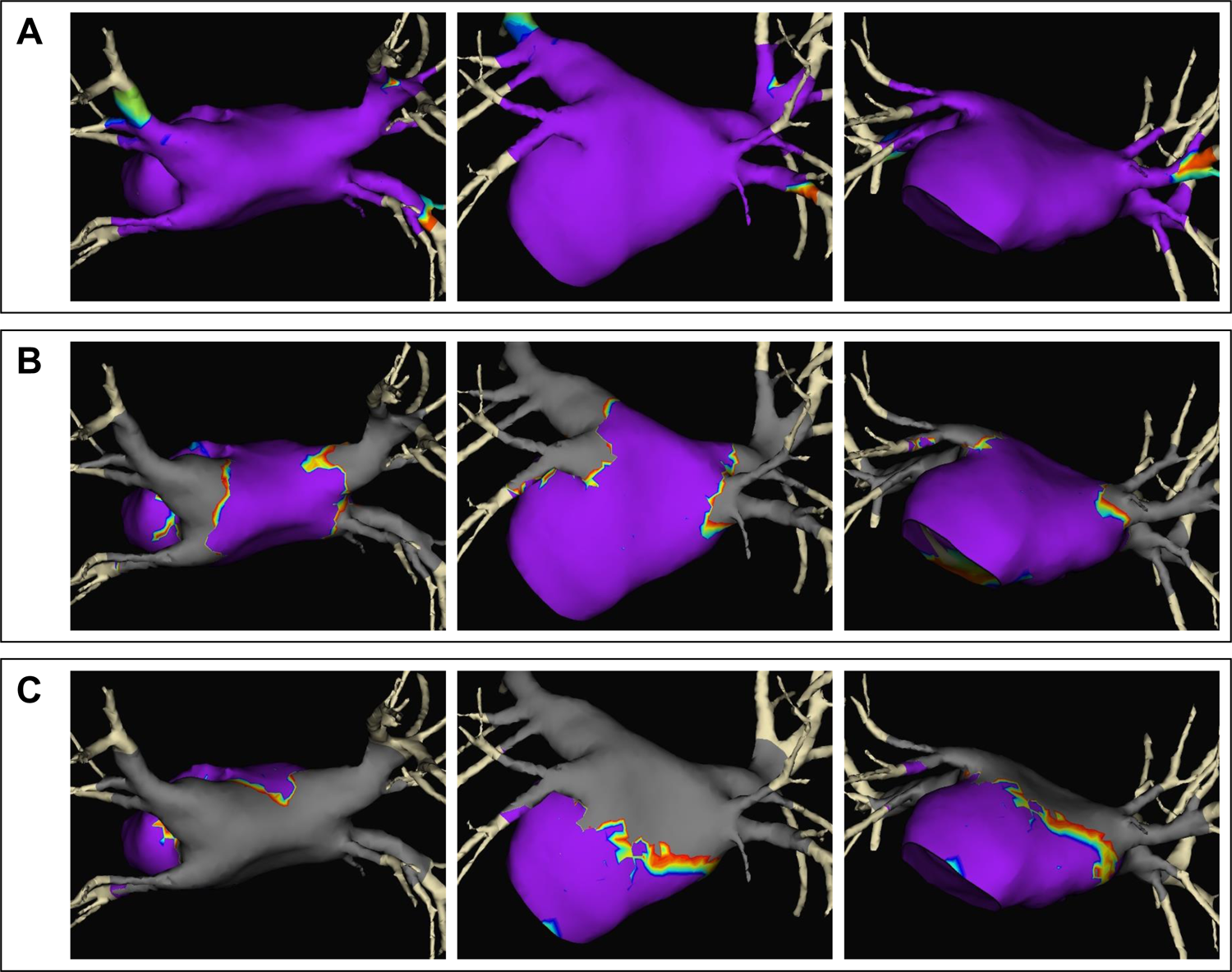
Representative voltage maps of the pre ablation (A), post PVI-only (B), and post PVI + LAPWI (C) recorded with the three-dimensional mapping with the EnSite Velocity system and Advisor^TM^ VL Circular Mapping Catheter (Abbott Laboratories, Chicago, IL, USA), voltage settings were 0.2 to 0.5mV. The purple region represented the voltages greater than 0.5mV and the gray region represented the voltages less than 0.2mV. PVI, pulmonary vein isolation; LAPWI, left atrial posterior wall isolation.

### Postprocedural management and Follow-up

After the procedure, patients were continuously monitored via telemetry until discharge, which was usually on the 2^nd^ postprocedural day. Previous prescriptions of AADs were resumed after the procedure at the discretion of the attending cardiologist. The patients were followed-up at our institution 1 month after ablation and every 2–3 months thereafter, or whenever patients experienced symptoms. A 12-lead electrocardiogram (ECG) was recorded at each visit to the outpatient clinic, and 24-h Holter ECG monitoring was performed at 3, 6, 12, and 24 months after the procedure. Recurrence was defined as any symptomatic or asymptomatic episode of atrial tachyarrhythmia (AF, atrial tachycardia [AT], or atrial flutter) lasting >30 s after the 3-month blanking period.

### Statistical analysis

Continuous data are expressed as the mean ± standard deviation for normally distributed variables or as the median with the interquartile range for non-normally distributed variables. Categorical variables are presented as numbers and percentages. Statistical comparisons were performed using Student’s t-test or Chi-square test, as appropriate. The analysis of the time-to-first event was described using Kaplan–Meier estimates; groups were compared using the log-rank test. All statistical analyses were performed using SPSS (version 29.0; IBM Company, Chicago, IL, USA), with *P* < 0.05 indicating statistical significance.

## Results

### Baseline characteristics

The baseline patient characteristics are shown in Table 1. In the PVI + PWI group, the mean age was 65.9 ± 10.5 years, 83 patients (76.9%) were male, and the median AF duration was (interquartile range: 3.0–10.0) months. There were no significant differences between the PVI + PWI and PVI-only groups in these parameters. The mean CHADS_2_ score, CHA_2_DS_2_-VASc score, and HAS-BLED score tended to be low in both groups. The mean left ventricular ejection fraction and mean LA diameter by echocardiography were 59.6 ± 11.3% and 43.5 ± 5.3mm, respectively, in the PVI + PWI group. There was no significant difference in each parameter between the PVI + PWI and PVI-only groups.

**Table 1.**
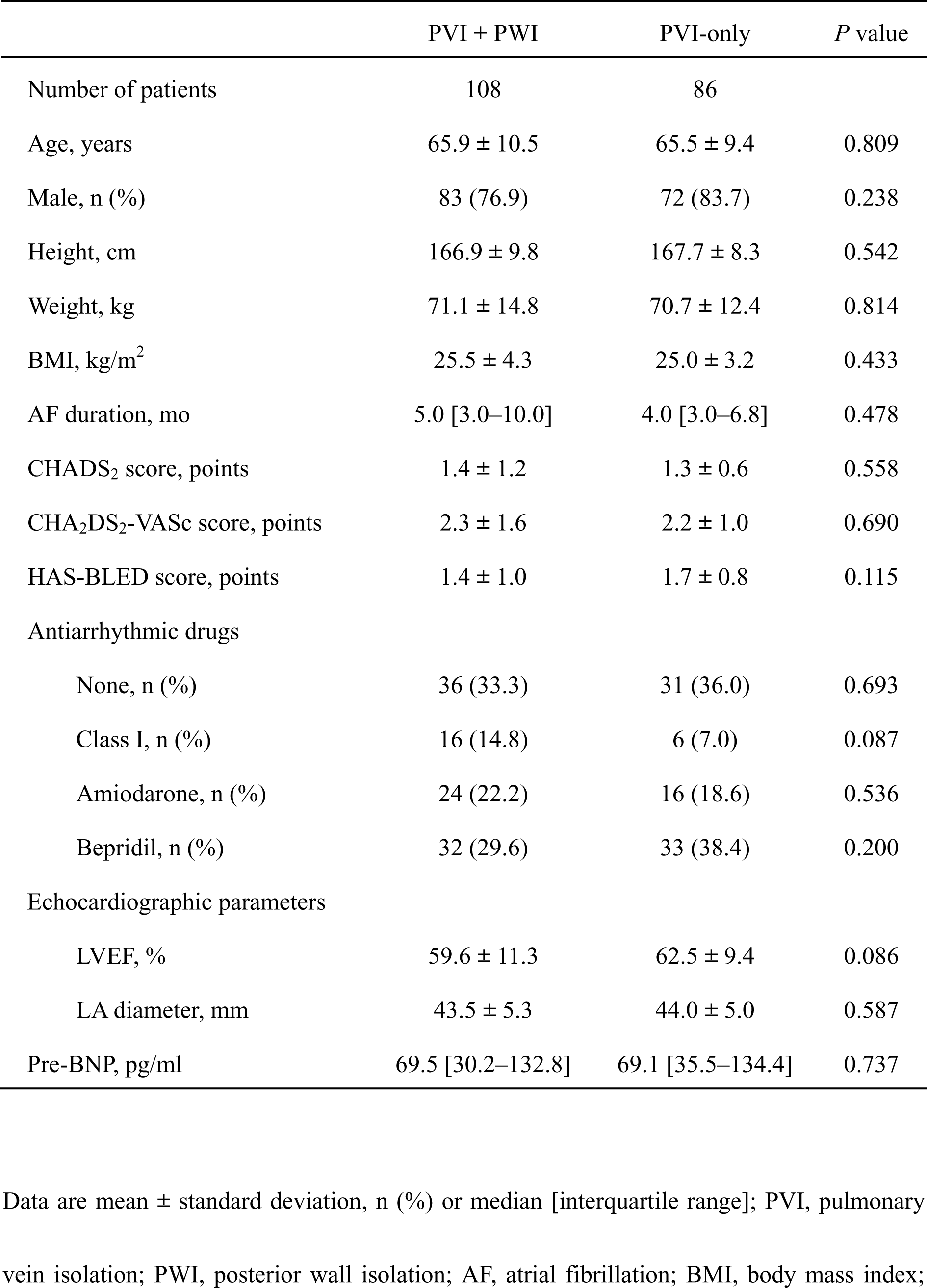
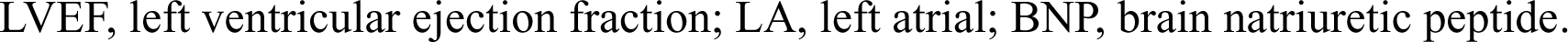
Baseline patient characteristic.

|  | PVI + PWI | PVI-only | *P* value |
| --- | --- | --- | --- |
| Number of patients | 108 | 86 |  |
| Age, years | 65.9 ± 10.5 | 65.5 ± 9.4 | 0.809 |
| Male, n (%) | 83 (76.9) | 72 (83.7) | 0.238 |
| Height, cm | 166.9 ± 9.8 | 167.7 ± 8.3 | 0.542 |
| Weight, kg | 71.1 ± 14.8 | 70.7 ± 12.4 | 0.814 |
| BMI, kg/m<sup>2</sup> | 25.5 ± 4.3 | 25.0 ± 3.2 | 0.433 |
| AF duration, mo | 5.0 [3.0–10.0] | 4.0 [3.0–6.8] | 0.478 |
| CHADS<sub>2</sub> score, points | 1.4 ± 1.2 | 1.3 ± 0.6 | 0.558 |
| CHA<sub>2</sub>DS<sub>2</sub>-VASc score, points | 2.3 ± 1.6 | 2.2 ± 1.0 | 0.690 |
| HAS-BLED score, points | 1.4 ± 1.0 | 1.7 ± 0.8 | 0.115 |
| Antiarrhythmic drugs |  |  |  |
| None, n (%) | 36 (33.3) | 31 (36.0) | 0.693 |
| Class I, n (%) | 16 (14.8) | 6 (7.0) | 0.087 |
| Amiodarone, n (%) | 24 (22.2) | 16 (18.6) | 0.536 |
| Bepridil, n (%) | 32 (29.6) | 33 (38.4) | 0.200 |
| Echocardiographic parameters |  |  |  |
| LVEF, % | 59.6 ± 11.3 | 62.5 ± 9.4 | 0.086 |
| LA diameter, mm | 43.5 ± 5.3 | 44.0 ± 5.0 | 0.587 |
| Pre-BNP, pg/ml | 69.5 [30.2–132.8] | 69.1 [35.5–134.4] | 0.737 |

Data are mean ± standard deviation, n (%) or median [interquartile range]; PVI, pulmonary vein isolation; PWI, posterior wall isolation; AF, atrial fibrillation; BMI, body mass index;
LVEF, left ventricular ejection fraction; LA, left atrial; BNP, brain natriuretic peptide.

### Procedural results

The procedural-related data are shown in Table 2. Acute PVI was achieved in all patients with CB without the need for touch-up RF ablation. The mean number of CB applications required for PVI was 5.2 ± 1.2, the mean total ablation time to achieve PVI was 14.9 ± 3.1 minutes, the mean minimum CB temperature reached to achieve PVI was −55.2℃ ± 3.7℃, and additional cavotricuspid isthmus (CTI) ablation was performed in 112 patients (57.7%), all of which had a confirmed CTI bidirectional block. The PVI + PWI group, compared to the PVI-only group, had significantly longer procedure time (180.0 ± 42.4 vs. 78.0 ± 22.1 min, *P* < 0.001) and fluoroscopy time (53.1 ± 17.4 vs. 21.3 ± 6.7 min, *P* < 0.001).

**Table 2.**
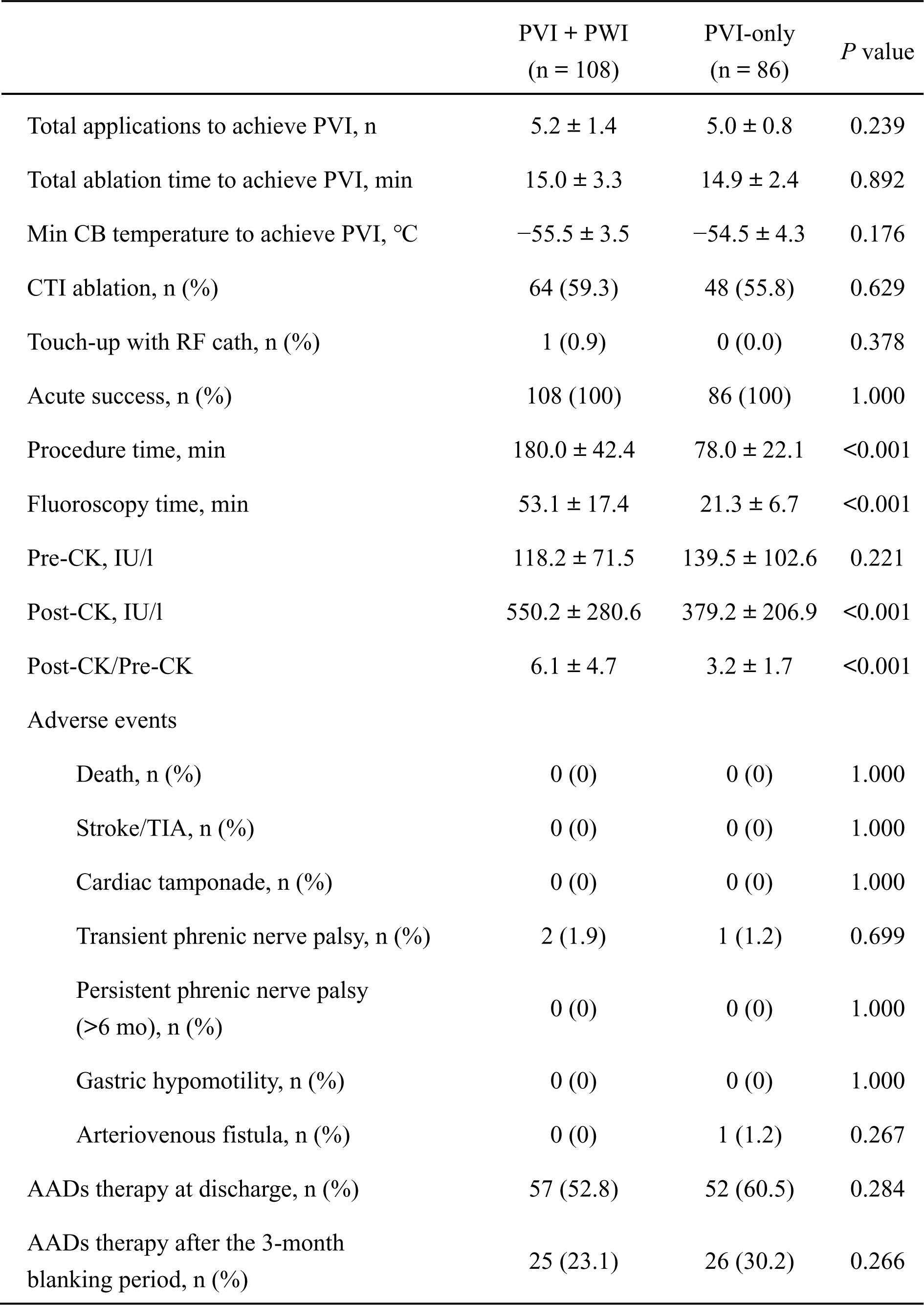
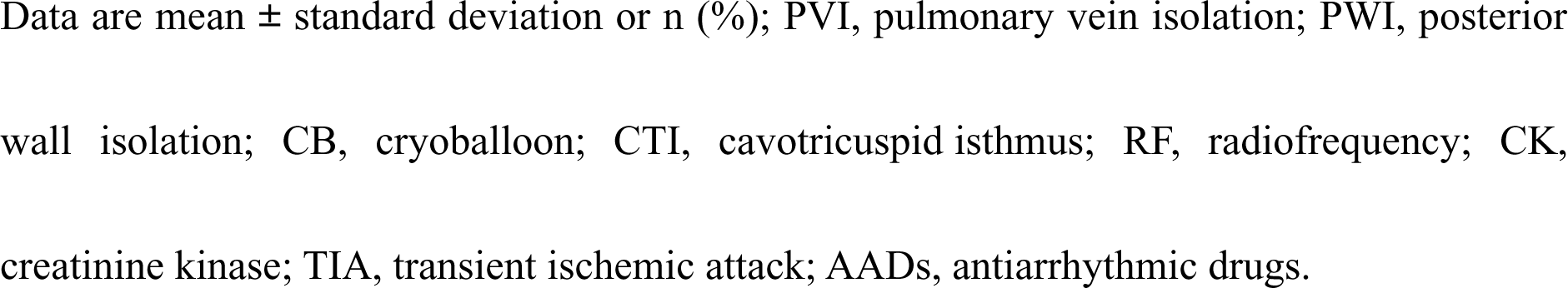
Procedural-related data and postprocedural AADs therapy.

|  | PVI + PWI<br>(n = 108) | PVI-only<br>(n = 86) | *P* value |
| --- | --- | --- | --- |
| Total applications to achieve PVI, n | 5.2 ± 1.4 | 5.0 ± 0.8 | 0.239 |
| Total ablation time to achieve PVI, min | 15.0 ± 3.3 | 14.9 ± 2.4 | 0.892 |
| Min CB temperature to achieve PVI, °C | -55.5 ± 3.5 | -54.5 ± 4.3 | 0.176 |
| CTI ablation, n (%) | 64 (59.3) | 48 (55.8) | 0.629 |
| Touch-up with RF cath, n (%) | 1 (0.9) | 0 (0.0) | 0.378 |
| Acute success, n (%) | 108 (100) | 86 (100) | 1.000 |
| Procedure time, min | 180.0 ± 42.4 | 78.0 ± 22.1 | <0.001 |
| Fluoroscopy time, min | 53.1 ± 17.4 | 21.3 ± 6.7 | <0.001 |
| Pre-CK, IU/l | 118.2 ± 71.5 | 139.5 ± 102.6 | 0.221 |
| Post-CK, IU/l | 550.2 ± 280.6 | 379.2 ± 206.9 | <0.001 |
| Post-CK/Pre-CK | 6.1 ± 4.7 | 3.2 ± 1.7 | <0.001 |
| Adverse events |  |  |  |
| Death, n (%) | 0 (0) | 0 (0) | 1.000 |
| Stroke/TIA, n (%) | 0 (0) | 0 (0) | 1.000 |
| Cardiac tamponade, n (%) | 0 (0) | 0 (0) | 1.000 |
| Transient phrenic nerve palsy, n (%) | 2 (1.9) | 1 (1.2) | 0.699 |
| Persistent phrenic nerve palsy<br>(>6 mo), n (%) | 0 (0) | 0 (0) | 1.000 |
| Gastric hypomotility, n (%) | 0 (0) | 0 (0) | 1.000 |
| Arteriovenous fistula, n (%) | 0 (0) | 1 (1.2) | 0.267 |
| AADs therapy at discharge, n (%) | 57 (52.8) | 52 (60.5) | 0.284 |
| AADs therapy after the 3-month<br>blanking period, n (%) | 25 (23.1) | 26 (30.2) | 0.266 |

Data are mean ± standard deviation or n (%); PVI, pulmonary vein isolation; PWI, posterior wall isolation; CB, cryoballoon; CTI, cavotricuspid isthmus; RF, radiofrequency; CK, creatinine kinase; TIA, transient ischemic attack; AADs, antiarrhythmic drugs.

LAPW was successfully isolated solely with the CB using the cross-over technique except for one case (99.1%). The level of serum creatine kinase (CK) the day after the procedure and the ratio of CK the day after to the day before the procedure were significantly higher in the PVI + PWI group than in the PVI-only group (550.2 ± 280.6 vs. 379.2 ± 206.9 IU/l, *P* < 0.001; 6.1 ± 4.7 vs. 3.2 ± 1.7, *P* < 0.001, respectively).

### LAPW isolation

The procedural characteristics of LAPWI are shown in Table 3. The mean number of CB applications was 8.6 ± 1.0 (L→R 4.3 ± 1.1 and R→L 4.3 ± 1.1) for LA roof ablation and 9.1 ± 1.2 (L→R 4.5 ± 1.2 and R→L 4.6 ± 1.6) for LA bottom ablation. The mean minimum CB temperature was −40.8℃ ± 5.4℃ (L→R −41.2℃ ± 5.7℃ and R→L −40.5℃ ± 4.7℃) during LA roof ablation (Figure 5A and 5C) and −37.8℃ ± 5.1℃ (L→R −37.9℃ ± 4.9℃ and R→L −37.7℃ ± 4.9℃) during LA bottom ablation (Figure 5B and 5D).

**Figure 5.**
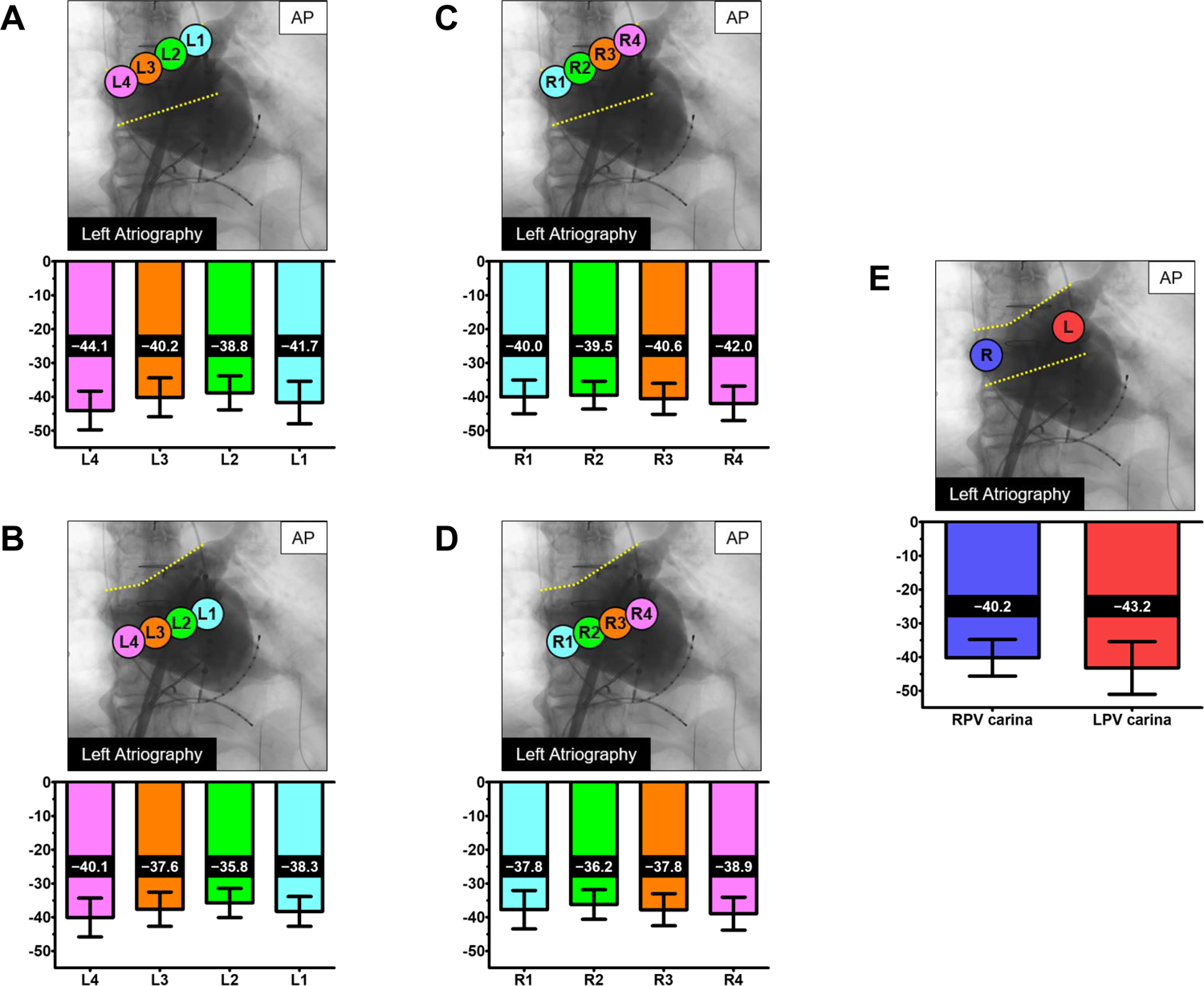
The mean minimum CB temperature at each site during the LA roof area (from left to right [A] and right to left [C] sides), LA bottom area (from left to right [B] and right to left [D] sides) and PV carinas (E) ablation. Data are mean ± standard deviation.

**Table 3.** Procedural characteristics of LAPWI.

|  | Applications, n | Minimum CB temperature, °C |
| --- | --- | --- |
| Roof, total | $8.6 \pm 1.0$ | $-40.8 \pm 5.4$ |
| Roof, L→R | $4.3 \pm 1.1$ | $-41.2 \pm 5.7$ |
| L1 | $1.0 \pm 0.1$ | $-41.7 \pm 6.3$ |
| L2 | $1.1 \pm 0.4$ | $-38.8 \pm 5.0$ |
| L3 | $1.1 \pm 0.4$ | $-40.2 \pm 5.7$ |
| L4 | $1.0 \pm 0.2$ | $-44.1 \pm 5.7$ |
| Roof, R→L | $4.3 \pm 1.1$ | $-40.5 \pm 4.7$ |
| R1 | $1.0 \pm 0.1$ | $-40.0 \pm 5.0$ |
| R2 | $1.0 \pm 0.2$ | $-39.5 \pm 4.1$ |
| R3 | $1.1 \pm 0.3$ | $-40.6 \pm 4.6$ |
| R4 | $1.2 \pm 0.4$ | $-42.0 \pm 5.1$ |
| Bottom, total | $9.1 \pm 1.2$ | $-37.8 \pm 5.1$ |
| Bottom, L→R | $4.5 \pm 1.2$ | $-37.9 \pm 4.9$ |
| L1 | $1.0 \pm 0.1$ | $-38.3 \pm 4.4$ |
| L2 | $1.3 \pm 0.5$ | $-35.8 \pm 4.3$ |
| L3 | $1.2 \pm 0.4$ | $-37.6 \pm 5.1$ |
| L4 | $1.0 \pm 0.1$ | $-40.1 \pm 5.7$ |
| Bottom, R→L | $4.6 \pm 1.6$ | $-37.7 \pm 4.9$ |
| R1 | $1.0 \pm 0.3$ | $-37.8 \pm 5.7$ |
| R2 | $1.1 \pm 0.4$ | $-36.2 \pm 4.4$ |
| R3 | $1.3 \pm 0.5$ | $-37.8 \pm 4.7$ |
| R4 | $1.2 \pm 0.4$ | $-38.9 \pm 4.9$ |
| LPV carina | $1.0 \pm 0.0$ | $-43.2 \pm 7.8$ |
| RPV carina | $1.0 \pm 0.0$ | $-40.2 \pm 5.4$ |
| LAPW (roof, bottom, and both PV carinas), total | $19.3 \pm 1.6$ | $-39.5 \pm 5.7$ |

Data are mean ± standard deviation or n (%); CB, cryoballoon; L→R, from left to right side; R→L, from right to left side; LPV, left pulmonary vein; RPV, right pulmonary vein; LAPW, left atrial posterior wall.

LPV and RPV carinas were ablated using CB in all patients in the PVI + PWI group. A mean number of 2.0 ± 0.0 CB applications (LPV 1.0 ± 0.0 and RPV 1.0 ± 0.0) was delivered for PV carina ablation. The mean minimum CB temperature during PV carina ablation was −41.7℃ ± 6.9℃ (LPV −43.2℃ ± 7.8℃ and RPV −40.2℃ ± 5.4℃) (Figure 5E). A mean number of 19.3 ± 1.6 CB applications was delivered for LAPW isolation (roof, bottom, and both PV carinas). The mean minimum CB temperature for total LAPW ablation was −39.5℃ ± 5.7℃ (Table 3). LAPW was successfully isolated solely using the CB in 107 out of 108 patients (99.1%), whereas the LAPW was completed by additional touch-up RF ablation in 1 patient (Table 2).

### Complications

The safety endpoints are shown in Table 2. The overall adverse event rate was low, with no significant difference between the PVI + PWI and PVI-only groups (1.9% vs. 2.3%, *P* = 0.818). Transient PN palsy occurred in 2 patients (1.9%) in the PVI + PWI group and 1 patient (1.2%) in the PVI-only group, with complete resolution within 6 months after the procedure. No PN palsy occurred during LA roof, bottom, or PV carina ablation. One patient (1.2%) in the PVI-only group developed an arteriovenous fistula which required surgical repair. No other severe periprocedural complications, such as death, stroke, cardiac tamponade, or GH, were observed.

### Follow-up

AADs were resumed in 109 out of 194 (56.2%) patients at discharge (PVI + PWI group, 52.8%; PVI-only group, 60.5%; *P* = 0.284) and maintained in 51 out of 194 (26.3%) patients after the 3-month blanking period (PVI + PWI group, 23.1%; PVI-only group, 30.2%; *P* = 0.266) (Table 2). During a median follow-up of 19 (interquartile range: 14–27) months, all enrolled patients completed the study protocol. During the follow-up period, freedom from recurrence of all atrial tachyarrhythmias was achieved in 93.5% (101/108) of the PVI + PWI group and 72.9% (69/86) of the PVI-only group (Figure 6A). Specifically, freedom from AF recurrence was seen in 96.3% (104/108) of the PVI + PWI group and 72.9% (69/86) of the PVI-only group (Figure 6B). On the other hand, freedom from AT recurrence was seen in 97.2% (105/108) of the PVI + PWI group and 100% (86/86) of the PVI-only group (Figure 6C). LAPWI was associated with a significantly lower rate of midterm recurrence of all atrial tachyarrhythmias (log-rank *P* = 0.011), particularly AF (log-rank *P* = 0.001), compared with PVI-only (Figure 6A and 6B).

**Figure 6.**
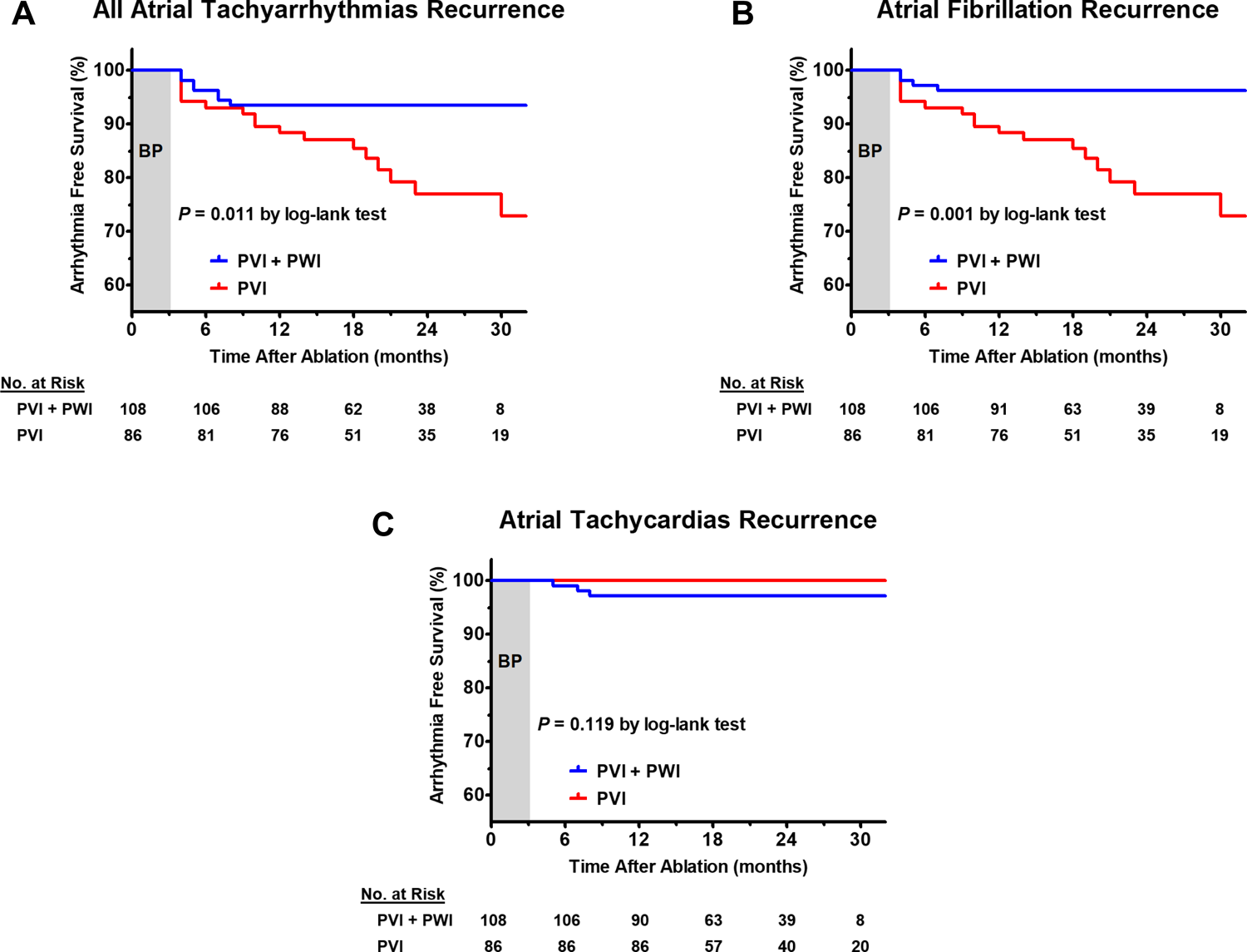
Kaplan-Meier Curves illustrating cumulative freedom from midterm recurrence of all atrial tachyarrhythmias (A), atrial fibrillation (B), and atrial tachycardias (C) after the procedure in the PVI + PWI group and in the PVI-only group, including the number of patients at risk. BP, blanking period.

Univariate and multivariate Cox regression analyses to identify the potential risk factors for the recurrence of all atrial tachyarrhythmias are shown in Table 4. After adjusting for confounding factors, LA diameter (hazard ratio [HR] 1.103, 95% confidence interval [CI] 1.005–1.211; *P* = 0.038) and PVI + PWI (HR 0.359, 95% CI 0.138–0.931; *P* = 0.035) were independent predictors of all atrial tachyarrhythmias recurrence.

**Table 4.**
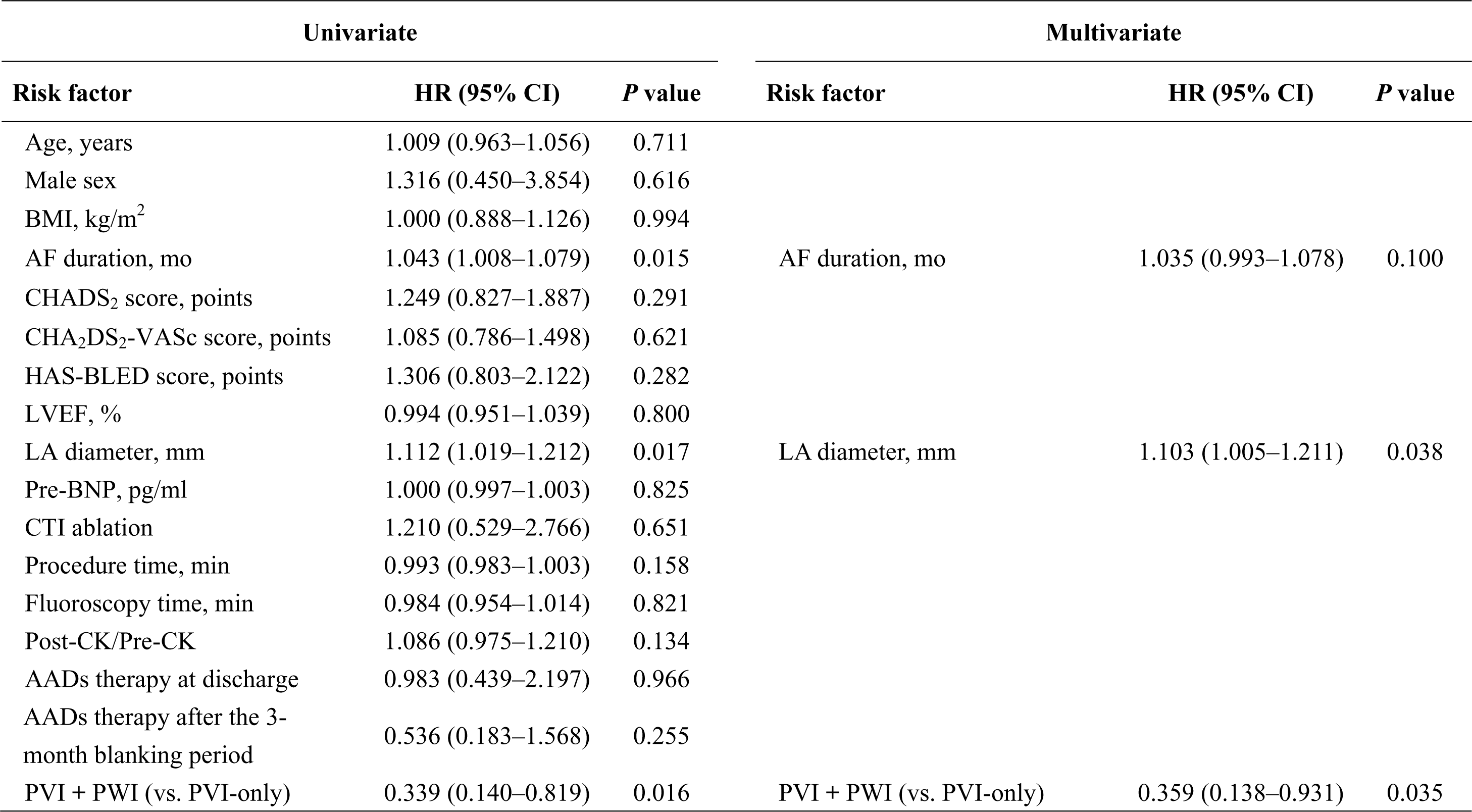
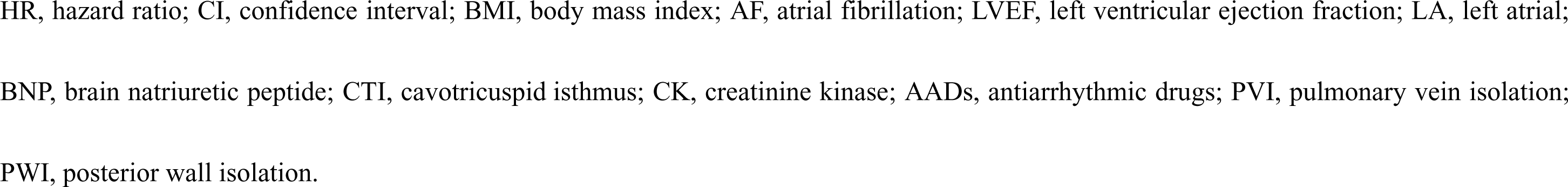
Predictors of all atrial tachyarrhythmias recurrence in a Cox regression analysis.

| Univariate |  |  | Multivariate |  |  |
| --- | --- | --- | --- | --- | --- |
| Risk factor | HR (95% CI) | P value | Risk factor | HR (95% CI) | P value |
| Age, years | 1.009 (0.963–1.056) | 0.711 |  |  |  |
| Male sex | 1.316 (0.450–3.854) | 0.616 |  |  |  |
| BMI, kg/m<sup>2</sup> | 1.000 (0.888–1.126) | 0.994 |  |  |  |
| AF duration, mo | 1.043 (1.008–1.079) | 0.015 | AF duration, mo | 1.035 (0.993–1.078) | 0.100 |
| CHADS<sub>2</sub> score, points | 1.249 (0.827–1.887) | 0.291 |  |  |  |
| CHA<sub>2</sub>DS<sub>2</sub>-VASc score, points | 1.085 (0.786–1.498) | 0.621 |  |  |  |
| HAS-BLED score, points | 1.306 (0.803–2.122) | 0.282 |  |  |  |
| LVEF, % | 0.994 (0.951–1.039) | 0.800 |  |  |  |
| LA diameter, mm | 1.112 (1.019–1.212) | 0.017 | LA diameter, mm | 1.103 (1.005–1.211) | 0.038 |
| Pre-BNP, pg/ml | 1.000 (0.997–1.003) | 0.825 |  |  |  |
| CTI ablation | 1.210 (0.529–2.766) | 0.651 |  |  |  |
| Procedure time, min | 0.993 (0.983–1.003) | 0.158 |  |  |  |
| Fluoroscopy time, min | 0.984 (0.954–1.014) | 0.821 |  |  |  |
| Post-CK/Pre-CK | 1.086 (0.975–1.210) | 0.134 |  |  |  |
| AADs therapy at discharge | 0.983 (0.439–2.197) | 0.966 |  |  |  |
| AADs therapy after the 3-month blanking period | 0.536 (0.183–1.568) | 0.255 |  |  |  |
| PVI + PWI (vs. PVI-only) | 0.339 (0.140–0.819) | 0.016 | PVI + PWI (vs. PVI-only) | 0.359 (0.138–0.931) | 0.035 |

HR, hazard ratio; CI, confidence interval; BMI, body mass index; AF, atrial fibrillation; LVEF, left ventricular ejection fraction; LA, left atrial; BNP, brain natriuretic peptide; CTI, cavotricuspid isthmus; CK, creatinine kinase; AADs, antiarrhythmic drugs; PVI, pulmonary vein isolation; PWI, posterior wall isolation.

## Discussion

Although the CB catheter is designed for single-shot PVI^20^, several studies have reported that PVI with concomitant LAPWI using the CB is superior to PVI alone for the treatment of PerAF ^12–16^, but no formal protocol has been established. To the best of our knowledge, this is the first study to describe the feasibility, safety, and efficacy of PVI with concomitant LAPWI solely with the CB using the cross-over technique. Our key findings are as follows: (1) complete LAPWI solely with the fourth-generation 28-mm CB catheter using the cross-over technique was achieved in 99.1% of PerAF patients; (2) PVI with concomitant LAPWI using the CB did not increase procedure-related complications; (3) the rate of midterm recurrence of atrial tachyarrhythmias was significantly lower in the PVI + PWI group than in the PVI-only group.

Nishimura et al. demonstrated that having an LA roof angle (i.e., the anatomical angle of the bucking point of the LA roof formed by the left-sided and right-sided roof) less than 115° predicted unsuccessful construction of an LA roof blockline because of the difficulty in ensuring optimal CB contact at the bucking point of a relatively narrow-angled LA roof^13^. Although this study had 6 patients (5.6%) with an LA roof angle < 115° in the PVI + PWI group, we were able to successfully construct an LA roof blockline solely with the CB using the cross-over technique in all patients of the PVI + PWI group. It was also difficult to ensure optimal CB contact with the concave and steep-angled LA bottom compressed by vertebral bodies. Nevertheless, we were able to successfully construct the LA bottom blockline in the PVI + PWI group solely with the CB using the cross-over technique, except for one case. The mean angle of the LA bottom was 140.0 ± 13.3° in the PVI + PWI group, whereas the unsuccessful case had the steepest angle of 103°. Thus, the cross-over technique could be considered an effective ablation strategy for difficult cases such as narrow and steeply angled LA roof and bottom, because this technique involves overlapping ablations by placing the CB from both left to right and right to left directions in contact with the roof and bottom of the LA.

Miyazaki et al^30^. reported that maneuverability for additional PWI might be superior with the fourth-generation versus the second-generation CB because of its relatively shorter tip length. Previous reports of LAPWI using the CB for PerAF have used the second-generation CB^12–16^. However, the fourth-generation CB was used in all patients in this study; its shorter balloon tip likely enabled optimal with the LA roof and bottom area, which might have also led to the high rates of acute success and freedom from atrial tachyarrhythmia recurrence.

Previous studies utilized a series of overlapping 120–240 s CB applications at each site during LAPWI^13–15^. In this study, 120-s CB applications were delivered at each site for LA roof and bottom ablation, while 180-s CB applications were delivered for PV carina ablation. Having a longer CB application time correlates with deeper lesions, leading to increased lesion durability^21^. However, it may also be associated with an increased risk of collateral damage to extracardiac structures, such as the PN and esophagus^22–24^. Esophageal thermal injury, which can lead to more serious conditions such as GH and left atrial-esophageal fistula, is a potential complication after LAPWI^25, 26^. Previous studies have reported that esophageal thermal injury rates associated with CB ablation ranged from 3.2% to 19% seen on postprocedural esophagogastroduodenoscopy (EGD)^27, 28^. In addition, Miyazaki et al. reported that the incidence of symptomatic GH was 0.23%^29^. In this study, no serious esophageal thermal injury such as symptomatic GH was observed, although mild esophageal lesions might have been missed because postprocedural EGD was not performed.

In areas where sufficient application time could not be secured due to the LET dropping below 18℃, CB application can often be achieved without reaching the LET of 18℃ or less by anchoring it from the opposite side, even if the balloon location is almost the same position under fluoroscopy. This is possible because the direction of force transmission is different when anchoring from the left and the right side; this is one of the benefits of the cross-over technique.

A single 120-s CB application might be insufficient for lesion formation and durability at each site. However, the cross-over technique allows for sequential overlapping freezes from both the left and right sides. This may have a beneficial impact on the efficacy and safety of LAPWI using the CB.

It is important not only to construct the horizontal block lines of the LA roof and bottom but also the vertical block lines consisting of the PV and PV carina to ensure durability of the LAPWI. Accordingly, LPV and RPV carinas were routinely ablated in addition to the 4 PVs, LA roof, and LA bottom in this study; this may have contributed to the high rate of freedom from atrial tachyarrhythmia recurrence.

A redo ablation procedure was performed in 3 out of 7 patients exhibiting atrial tachyarrhythmia recurrence in the PVI + PWI group. Two patients had a recurrence of AF and the other patient had a recurrence of AT. The patients with AF recurrence were confirmed to have reconnection at the LSPV (and CTI) and RIPV, respectively, and the patient with AT recurrence was confirmed at the perimitral AT. These three patients were also confirmed to have no reconnection at the LA roof, LA bottom, and both PV carinas. On the other hand, Shigeta et al^14^. reported that LA roof and bottom-line ablation performed using the CB demonstrated durability in 72.5% and 48.3% of patients, respectively, at the beginning of the redo ablation procedure.

The lower durability of LA bottom ablation versus that of LA roof ablation using the CB is due to the difficulty in ensuring the optimal contact of the distal hemisphere of the CB to the LA bottom. In addition, inadequate CB contact due to the steep-angled LA bottom and insufficient application time due to reaching the LET cut-off value might also be contributory. The cross-over technique for LAPWI using the CB could be effective against any of these challenging factors. Although further studies are required to evaluate the durability of the LAPWI performed with the CB using the cross-over technique, it can potentially improve the acute success rate and durability of LAPWI.

The number of CB applications in the PVI + PWI group led to significantly longer procedure and fluoroscopy times and significantly higher postprocedural CK levels versus the PVI-only group. However, no severe adverse events associated with these factors (e.g., radiodermatitis and pericarditis) were observed.

## Limitations

Several limitations of this study should be acknowledged. First, this was a single-center, non-randomized, retrospective, observational study with a relatively small study population. Even if all variables were balanced between the two groups, selection bias cannot be excluded. Second, esophageal damage might have been underestimated because EGD was not performed after the procedure. Third, despite the absence of a significant difference in AAD therapy between the two groups, this was still maintained in 26.3% of patients after the 3-month blanking period, which may have influenced our findings. Fourth, the rate of symptomatic and asymptomatic atrial tachyarrhythmia recurrence might have been underestimated, because no long-term monitoring was done (i.e., using an event recorder). Lastly, all procedures were performed by a single operator who was experienced in the cross-over technique, and thus these results may be dependent on the skill of the operator. Further studies, including multicenter and large populations, are needed to confirm the general usefulness of this ablation technique.

## Conclusions

Despite having significantly longer procedure and fluoroscopy times, LAPWI performed solely with the CB using the cross-over technique is feasibly, safe, and independently associated with a significantly higher freedom from recurrence of atrial tachyarrhythmias compared with PVI alone on midterm follow-up in patients with PerAF. This technique can potentially improve the acute success rate and durability of LAPWI.

## Data Availability

Raw data were generated at Juntendo University Urayasu Hospital. Derived data supporting the findings of this study are available from the corresponding author [FO] on request.

## Non-standard Abbreviations and Acronyms

CB: cryoballoon
PVI: pulmonary vein isolation
LAPWI: left atrial posterior wall isolation
LA: left atrium
PerAF: persistent atrial fibrillation
OACs: oral anticoagulants
TTE: transthoracic echocardiogram
AADs: antiarrhythmic drugs
GH: gastric hypomotility
LET: luminal esophageal temperature
PN: phrenic nerve
CMAP: compound motor action potentials
RF: radiofrequency
AP: anteroposterior
ECG: electrocardiogram
AT: atrial tachycardia
CTI: cavotricuspid isthmus
CK: creatine kinase
EGD: esophagogastroduodenoscopy

